# Home-based primary care services for managing non-communicable diseases in rural India: a retrospective multiple time-point observational study

**DOI:** 10.64898/2026.10.28.26366753

**Authors:** Uma Gupta, Lakshmi Krishnan, Shreyas Patil, Jashanjot Singh Mangat, Richa Shete, Siddhesh Zadey

## Abstract

**Purpose:** Limited access to healthcare can worsen the burden of non-communicable diseases (NCDs). Home-based primary care models, supported by trained health workers enabled with digital platforms, offer a potential strategy to improve NCD prevention and management. Yet, evidence on how such models perform in real-world settings is limited.

**Methods:** We conducted a retrospective analysis of routinely collected data from September 2022 to March 2025 on participants in NEEM, a subscription-based, home-delivered primary care program for managing NCDs in rural Maharashtra, India. Participants received regular home visits for screening, monitoring, medication delivery, and telemedicine follow-up. Among participants with >1 visit, >30 days of follow-up, and at least two complete observations, we used generalized estimating equations to estimate within-person changes in systolic blood pressure (SBP), diastolic blood pressure (DBP), and random blood sugar (RBS), overall and by control status at screening, and changes in the population-average odds of hypertension (HTN) and diabetes (DM) control.

**Results:** The primary analytic sample comprised 1,668 participants (12,769 observations; median follow-up 252.5 days). SBP changed by −1.19 mmHg (95% CI: −1.38, −1.01), DBP by −0.75 mmHg (−0.86, −0.64), and RBS by −0.77 mg/dL (−1.40, −0.14) per 90 days. The odds of HTN control increased by 21% (adjusted odds ratio [aOR]: 1.21 [95% CI: 1.17, 1.25]) and of DM control by 6% (aOR: 1.06 [1.04, 1.08]) per 90 days.

**Conclusions:** Among NEEM-engaged participants, blood pressure improved, with weaker evidence for blood sugar. Comparative evaluations are needed to establish whether such models improve control relative to traditional facility-based care.

## 1 Introduction

Non-communicable diseases (NCDs) are chronic conditions with complex sequelae that require multiple treatments and long-term care. The Global Burden of Disease study estimates that the share of disability-adjusted life-years attributable to NCDs has increased from about 41.7% to 64.4% between 1990 and 2023, underscoring the need to prioritize NCDs in global public health efforts.^1^ Uncontrolled hypertension (HTN) can lead to complications like myocardial infarction and stroke, and uncontrolled type 2 diabetes mellitus (DM) can lead to organ dysfunction, particularly affecting the kidneys, nerves, eyes, heart, and blood vessels over time.^2,3^ Initiatives like the WHO HEARTS technical package and the WHO Package of Essential Noncommunicable Disease Interventions for Primary Health Care (WHO PEN) list activities that can be easily deployed through locally recruited health workers.^4,5^

Given India’s large population and the demographic and epidemiologic transitions, the burden of NCDs is particularly high and continues to rise.^6^ As of 2021, an estimated 315 million people lived with HTN, and 101 million with DM.^6^ The National NCD Monitoring Survey 2017–18 reported that, among adults with HTN, only 29.2% were aware of their HTN status, 16% were receiving treatment, and only 12.3% had achieved adequate control.^7^ The National Family Health Survey (NFHS-5) 2019–21 estimated that among people with DM, 27.5% were aware of their condition, 21.5% were taking treatment, and only 7.0% (6.8–7.1%) had achieved glycemic control, with a significant difference between the highest and the lowest wealth quintiles.^8^

Recognizing the growing burden of disease, the Ministry of Health and Family Welfare, Government of India, launched the National Programme for Prevention and Control of NCDs, which aims to strengthen the continuum of care through screening, detection, management, referral, and follow-up at all levels of healthcare.^9^ It also focuses on overarching activities such as health promotion, capacity-building, strengthening supply-chain management, and intersectoral coordination. However, implementation has been hampered by factors including delays in symptom detection and diagnosis, as well as limited access to healthcare providers and medicines.^10^ The India Hypertension Control Initiative (IHCI), launched in 2017 to strengthen HTN management in public primary care settings and built on the WHO HEARTS initiative, showed that a scalable public health control program can drive significant improvements.^11–13^ Community health worker-led NCD programs in India with home components have mainly included screening and risk assessment, with diagnosis and treatment at primary health centers.^14,15^ Evidence on clinical management of HTN and DM delivered at home, including prescription and medication delivery and long-term evaluation, remains limited.

In private-sector primary care settings, using the protocols set by the WHO HEARTS initiative and IHCI for HTN and WHO PEN for DM, Dvara Health Finance (DHF) conceptualized a service model called NEEM aimed at households characterized as the “missing middle”, i.e., those not eligible for government-funded health insurance schemes and not currently covered by private health insurance.^16^ NEEM offers bundled healthcare services, including access to the care team and medication for HTN and DM in a monthly subscription format. The care team includes locally recruited health workers, remote doctors, and specialists.

Health workers conduct home visits to screen patients and record findings in the platform’s mobile application. If the application indicates it, patients connect with a remote doctor for further management. Additionally, health workers facilitate medicine orders and delivery at patients’ request. To ensure program adherence, as noted in IHCI, staff follow up with patients through monthly home visits to measure vital signs and provide medication and lifestyle counseling.^11–13^ Where hospitalization or advanced diagnostics are needed, the care team facilitates referral to a local network of partner hospitals. The program currently operates in three blocks of the Satara district in Maharashtra, India.

DHF’s in-house clinical decision-support platform, NEEM360, supports these services. The platform dynamically classifies patients into straight-through or referral pathways and provides clear, specific guidance to local health workers.^17^ The management of patients with no comorbidities or complex medical history is guided by evidence-based WHO HEARTS and PEN protocols. The platform generates a draft prescription in accordance with these protocols, after which a remote doctor connects with the patient via telemedicine. The doctor confirms the diagnosis, following which the prescription is sent immediately to the patient’s phone, and generic medicines are delivered within 3–4 days as part of the subscription. Individuals already on medications or those with comorbidities requiring tailored medical management are referred to specialists.

In this study, we assessed how participants engaging with home-based primary care through the NEEM program managed their HTN and DM outcomes. We used systolic and diastolic blood pressures (SBP and DBP) for HTN and random blood sugar (RBS) for DM outcomes. Specifically, we investigated the following questions for the engaged participants: (1) What is the within-person change in SBP, DBP, and RBS over follow-up? (2) How do the odds of HTN and DM control change over follow-up? (3) As an exploratory question, among those with uncontrolled HTN or DM at screening, which characteristics are associated with time to first observed control?

## 2 Methods

### 2.1 Study setting

The study setting spanned three blocks in Satara district, Maharashtra **(Supplementary Figure 1)**. In Maharashtra, NFHS-5 (2019–21) found that only about 11% of individuals with HTN had their blood pressure under control.^18,19^ Similarly, the Longitudinal Ageing Study in India Wave 1 (2017–18) reported that only 23% of adults aged 45 years and older with HTN and 28% of those with DM in the state achieved disease control.^20^ NFHS-5 also estimates HTN prevalence at approximately 24% in Satara district, though district-level data on control remain limited.^19^

### 2.2 Dataset and variables

We conducted a retrospective analysis of data collected under the NEEM program from September 22, 2022, to March 12, 2025. Trained health workers obtained informed consent from participants at the initial screening visit for data collection and use. DHF anonymized the data before sharing with the study investigators. This study was approved by the Indian Association for Preventive and Social Medicine Ethics Committee (IAPSM/00112024).

At baseline (i.e., screening), DHF collected sociodemographic details, anthropometric measures, and clinical parameters. Sociodemographic variables included age, sex, household/family size, and village of residence, as well as program engagement (active user status) and enrollment package type (advanced vs. base, depending on the services provided under the subscription). Anthropometric measures included height, weight, body mass index (BMI), and waist circumference. We categorized BMI as underweight (<18.5 kg/m²), normal (18.5–22.9 kg/m²), overweight (23.0–24.9 kg/m²), or obese (≥25.0 kg/m²) using Asia-Pacific cut-offs.^21^ Clinical/vital parameters included SBP, DBP, RBS, oxygen saturation (SpO□), pulse rate, respiratory rate, and body temperature. Health workers also recorded behavioral and clinical history, including smoking status; symptom and condition history such as chest pain and history of acute asthma; pre-existing conditions including chronic kidney disease and cardiovascular disease; and use of HTN and DM medications. We had multiple time-point data on SBP, DBP, and RBS, with unequal follow-up lengths and unequal between-observation intervals across participants.

### 2.3 Analytic dataset

For longitudinal analyses, we restricted the dataset to participants with evidence of continued engagement in the program. The merged program data consisted of 18,529 observations from 4,703 participants. We included participants with more than one recorded visit and more than 30 days from screening to the latest visit, set physiologically implausible values to missing **(Supplementary Table 1)**, and required complete data on the model variables and at least two complete observations per participant. The final primary analytic sample comprised 12,769 observations from 1,668 participants **(Figure 1)**.

**Figure 1:**
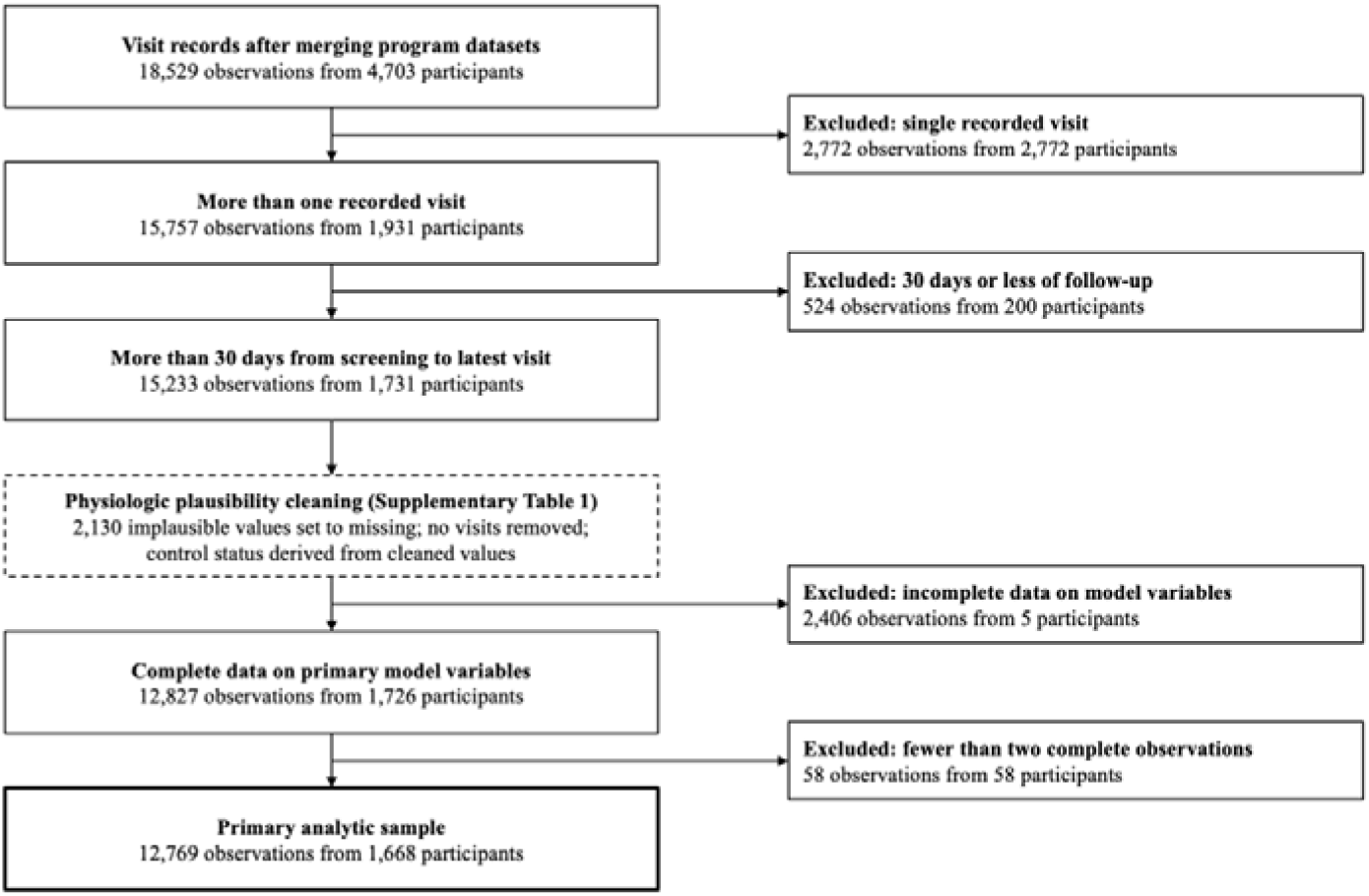
Derivation of the primary analytic sample.

For time-to-control analyses, we constructed a separate person-level dataset for each control outcome, restricted to participants uncontrolled for that outcome at screening. To avoid conditioning on more than 30 days of future follow-up, we drew these participants from those with more than one recorded visit, before applying the 30-day follow-up restriction, and then applied the same plausibility and complete-case steps. Each participant contributed one record: the time from screening to the first follow-up visit at which control was observed, or to the last observed follow-up measurement if control was not observed.

### 2.4 Outcomes

We defined HTN control as SBP <140 mmHg and DBP <90 mmHg.^22^ We also analyzed SBP control (SBP <140 mmHg) and DBP control (DBP <90 mmHg) separately. We defined DM control as RBS <200 mg/dL.^23^ No diagnosis variable was available, so we defined control from measured values in all screened participants, irrespective of prior diagnosis.

The primary outcomes were continuous values of SBP, DBP, and RBS measured at repeated follow-up visits, analyzed as within-person change over time. Secondary outcomes were HTN, SBP, DBP, and DM control at each follow-up visit. In exploratory analyses, we examined the time from screening to the first follow-up visit at which control was observed, among participants uncontrolled at screening. We measured these outcomes through serial follow-up records collected during routine program implementation. We reported control at the latest visit descriptively.

### 2.5 Statistical analysis

For the first research question, we evaluated longitudinal changes in SBP, DBP, and RBS using generalized estimating equations (GEE) with an identity link and Gaussian variance, an exchangeable working correlation structure, and robust (sandwich) standard errors. We measured time in days from screening to each follow-up visit, using the same time scale for all three outcomes. For each outcome, we first fit an unadjusted model, and then a multivariable model that included baseline covariates with complete data, selected a priori: age, sex, smoking history, known chronic kidney disease, known cardiovascular disease, and control of the other condition at screening (DM control in SBP and DBP models; HTN control in RBS models). We did not adjust for program user status, because it reflects ongoing engagement rather than a fixed baseline characteristic.

In this approach, a population-average GEE time coefficient can conflate genuine within-person change with between-person differences in composition along follow-up. Hence, we separated follow-up time into each participant’s mean follow-up time and the within-person deviation from that mean, and took the coefficient on the within-person deviation as the primary estimate of change (within-between, or Mundlak, decomposition).^24,25^ We reported the corresponding population-average slope as a sensitivity analysis. We did not adjust for the baseline value of the outcome, to avoid reintroducing regression to the mean. Under heterogeneous individual slopes, the within-person estimate is a weighted average that gives more weight to participants with more numerous and more widely spaced visits. The modeled series comprised follow-up visit records, including any recorded on the screening date; the screening measurement defined baseline control status and covariates. We reported change per 90 days, which more closely matches the observed follow-up, and annual change as a linear projection. To assess heterogeneity by baseline control, we fit adjusted models stratified by screening control status (HTN control for SBP and DBP; DM control for RBS) and models with time-by-screening-control interaction terms.

For the second research question, we fit logistic GEE models for HTN, SBP, DBP, and DM control at each follow-up visit, with the same working correlation, standard errors, and adjustment sets as noted above. For these models, the population-average estimate was primary, because the within-between decomposition is only approximate in logistic models; in the exploratory joint control-group models, we also reported a within-person version that added each participant’s mean follow-up time.

We quantified regression to the mean in each screening-control stratum by comparing the observed change from screening to the latest visit with the change expected from mean reversion, approximated as (r − 1) × (stratum mean at screening − overall mean at screening), where r is the correlation between participants’ screening and latest values **(Supplementary Table 2)**. We compared within-person and population-average slopes **(Supplementary Table 3)** and repeated the adjusted population-average models in participants with more than 90 days of follow-up **(Supplementary Table 4)**. In exploratory analyses, we cross-classified participants by joint HTN and DM control at screening and fit models with time interacted with the four-group variable, omitting the screening control covariates **(Supplementary Tables 5–6)**. We tested whether time slopes differed across the four groups with a joint Wald test.

For the third exploratory research question on time to control among participants uncontrolled at screening, we fit Cox proportional hazards models for the time from screening to the first follow-up visit at which HTN, SBP, DBP, or DM control was observed. Participants without observed control were censored at their last follow-up, and models were adjusted for the same covariates as the GEE models. Because the primary analytic sample required more than 30 days of follow-up, using it with measurement starting at screening could introduce immortal-time bias. We drew the analytic sample for the exploratory time-to-control analysis before the 30-day restriction as described in **Section 2.3**. We assessed the proportional hazards assumption with Schoenfeld residual tests. Because control could be observed only at visits, event times were interval-censored, and no event or censoring could occur before a participant’s first follow-up visit. However, we did not model these features separately. We used an alpha threshold of 0.05 for statistical significance.

We conducted all analyses and visualizations in R (Version 4.4.3) and validated and annotated code using Claude Opus 5.5 (Cowork).

## 3 Results

### 3.1 Descriptive statistics

The primary analytic sample comprised 1,668 participants who contributed 12,769 visit-level observations (2–25 per participant). Median follow-up from screening to the latest visit was 252.5 days (interquartile range 99–485), and 37% of participants were followed for more than one year.

At screening, participants had a mean age of 55.9 years (SD 13.3), and 912 (55%) were female **(Table 1)**. Overall, 924 (55%) participants were obese. HTN was uncontrolled in 719 (43%) participants, and DM in 452 (27%). At screening, 196 (12%) participants had both conditions uncontrolled, 256 (15%) had controlled HTN and uncontrolled DM, 523 (31%) had uncontrolled HTN and controlled DM, and 693 (42%) had both HTN and DM controlled. Mean SBP was 137.4 mmHg (SD 20.8), mean DBP 85.2 mmHg (SD 12.5), and mean RBS 170.0 mg/dL (SD 95.4). At the latest visit, HTN was controlled in 1,292 (77%) of 1,668 participants and DM in 1,185 (77%) of 1,547 participants with a recorded RBS.

**Table 1:** Characteristics of the primary analytic sample at screening and at the latest visit (N=1,668).

| **Characteristic** | **N=1,668** |
| --- | --- |
| SBP at screening, mmHg | 137.44 (20.82) |
| DBP at screening, mmHg | 85.22 (12.48) |
| RBS at screening, mg/dL | 170.02 (95.36) |
| **HTN control at screening** |  |
| Controlled | 949 (57%) |
| Uncontrolled | 719 (43%) |
| **DM control at screening** |  |
| Controlled | 1,216 (73%) |
| Uncontrolled | 452 (27%) |
| SBP at latest visit, mmHg | 130.45 (14.32) |
| DBP at latest visit, mmHg | 81.35 (9.45) |
| RBS at latest visit, mg/dL | 160.72 (76.28) |
| Missing | 121 |
| **HTN control at latest visit** |  |
| Controlled | 1,292 (77%) |
| Uncontrolled | 376 (23%) |
| **DM control at latest visit** |  |
| Controlled | 1,185 (77%) |
| Uncontrolled | 362 (23%) |
| Missing | 121 |
| Follow-up duration, days | 317.16 (246.24) |
| Age, years | 55.88 (13.30) |
| **Sex** |  |
| Female | 912 (55%) |
| Male | 756 (45%) |
| **Family size** |  |
| 1–2 | 198 (54%) |
| 3–4 | 151 (41%) |
| ≥5 | 17 (4.6%) |
| Missing | 1,302 |
| **Smoking history** |  |
| Current smoker | 84 (5.0%) |
| Former smoker | 21 (1.3%) |
| Non-smoker | 1,563 (94%) |
| **Body mass index** |  |
| Underweight | 84 (5.0%) |
| Normal | 363 (22%) |
| Overweight | 297 (18%) |
| Obese | 924 (55%) |
| Known chronic kidney disease | 17 (1.0%) |
| Known cardiovascular disease | 123 (7.4%) |
| DM medication use | 514 (68%) |
| Missing | 910 |
| HTN medication use | 497 (45%) |
| Missing | 564 |
| **Program user status** |  |
| Active | 918 (55%) |
| Inactive | 750 (45%) |
| **Package type** |  |
| Base | 965 (58%) |
| Advanced | 702 (42%) |
| Missing | 1 |

Data are mean (SD) or n (%); percentages are of participants with non-missing data. HTN control: SBP <140 mmHg and DBP <90 mmHg. DM control: RBS <200 mg/dL. Body mass index categories use Asia-Pacific cut-offs. SD: standard deviation; HTN: hypertension; DM: diabetes mellitus; SBP: systolic blood pressure; DBP: diastolic blood pressure; RBS: random blood sugar.

### 3.2 Within-person changes in SBP, DBP, and RBS

In the primary analytic sample, SBP changed by −1.19 mmHg per 90 days (95% CI: −1.38, −1.01), DBP by −0.75 mmHg per 90 days (−0.86, −0.64), and RBS by −0.77 mg/dL per 90 days (−1.40, −0.14) **(Table 2)**. Projected over one year, these correspond to reductions of 4.84 mmHg, 3.04 mmHg, and 3.12 mg/dL, respectively. Unadjusted and adjusted within-person estimates were identical because all adjustment covariates were fixed at baseline.

**Table 2:** Within-person change in SBP, DBP, and RBS over follow-up.

| **Model** | **Population (N)** | **Daily**<br>**coefficient**<br>**(SE)** | **Change per**<br>**90 days (95%**<br>**CI)** | **Projected**<br>**annual change**<br>**(95% CI)** | **P**<br>**value** |
| --- | --- | --- | --- | --- | --- |
| **SBP, mmHg** |  |  |  |  |  |
| Model 1:<br>unadjusted | Primary analytic<br>sample (1,668) | -0.0133<br>(0.0010) | -1.19 (-1.38,<br>-1.01) | -4.84 (-5.58,<br>-4.10) | <0.001 |
| Model 2:<br>adjusted |  | -0.0133<br>(0.0010) | -1.19 (-1.38,<br>-1.01) | -4.84 (-5.58,<br>-4.10) | <0.001 |
| Model 3:<br>adjusted | Uncontrolled<br>HTN at<br>screening (719) | -0.0270<br>(0.0016) | -2.43 (-2.71,<br>-2.15) | -9.86 (-10.98,<br>-8.74) | <0.001 |
| Model 4:<br>adjusted | Controlled HTN<br>at screening<br>(949) | 0.0011<br>(0.0009) | 0.10 (-0.06,<br>0.26) | 0.41 (-0.25,<br>1.06) | 0.224 |
| **DBP, mmHg** |  |  |  |  |  |
| Model 1:<br>unadjusted | Primary analytic<br>sample (1,668) | -0.0083<br>(0.0006) | -0.75 (-0.86,<br>-0.64) | -3.04 (-3.49,<br>-2.58) | <0.001 |
| Model 2:<br>adjusted |  | -0.0083<br>(0.0006) | -0.75 (-0.86,<br>-0.64) | -3.04 (-3.49,<br>-2.58) | <0.001 |
| Model 3:<br>adjusted | Uncontrolled<br>HTN at<br>screening (719) | -0.0157<br>(0.0010) | -1.41 (-1.59,<br>-1.24) | -5.73 (-6.44,<br>-5.02) | <0.001 |
| Model 4:<br>adjusted | Controlled HTN<br>at screening<br>(949) | -0.0006<br>(0.0006) | -0.05 (-0.16,<br>0.06) | -0.21 (-0.66,<br>0.24) | 0.350 |
| **RBS, mg/dL** |  |  |  |  |  |
| Model 1:<br>unadjusted | Primary analytic<br>sample (1,668) | -0.0085<br>(0.0036) | -0.77 (-1.40,<br>-0.14) | -3.12 (-5.66,<br>-0.57) | 0.016 |
| Model 2:<br>adjusted |  | -0.0085<br>(0.0036) | -0.77 (-1.40,<br>-0.14) | -3.12 (-5.66,<br>-0.57) | 0.016 |
| Model 3:<br>adjusted | Uncontrolled<br>DM at screening | -0.0842<br>(0.0115) | -7.57 (-9.59,<br>-5.55) | -30.71 (-38.91,<br>-22.52) | <0.001 |
|  | (452) |  |  |  |  |
| Model 4:<br>adjusted | Controlled DM<br>at screening<br>(1,216) | 0.0148<br>(0.0025) | 1.33 (0.89,<br>1.78) | 5.41 (3.61,<br>7.21) | <0.001 |

Among participants with uncontrolled HTN at screening (n=719), SBP changed by −2.43 mmHg per 90 days (−2.71, −2.15) and DBP by −1.41 mmHg per 90 days (−1.59, −1.24). Among those controlled at screening (n=949), neither SBP (0.10 mmHg per 90 days; 95% CI: −0.06, 0.26) nor DBP (−0.05; −0.16, 0.06) changed significantly. Among participants with uncontrolled DM at screening (n=452), RBS changed by −7.57 mg/dL per 90 days (−9.59, −5.55), whereas among those controlled at screening (n=1,216), RBS increased by 1.33 mg/dL per 90 days (0.89, 1.78). Slopes differed by screening control status for all three outcomes (interaction p<0.001); per 90 days, the slope among participants controlled at screening was 2.53 mmHg higher for SBP, 1.36 mmHg higher for DBP, and 8.91 mg/dL higher for RBS than among those uncontrolled.

Regression to the mean contributed to these stratified trajectories **(Supplementary Table 2)**. Between screening and the latest visit, mean SBP fell by 18.96 mmHg among participants with uncontrolled HTN at screening, compared with a 10.16 mmHg fall expected from regression to the mean. Among those with controlled HTN, mean SBP rose by 2.08 mmHg, compared with an expected rise of 7.70 mmHg. Findings for DBP showed a similar pattern. Among participants with uncontrolled DM at screening, mean RBS fell by 76.10 mg/dL, compared with an expected fall of 51.78 mg/dL. Among those with controlled DM, by contrast, RBS rose by 11.66 mg/dL, compared with an expected rise of 21.31 mg/dL.

Population-average slopes were similar to within-person slopes for SBP (−1.06 vs −1.19 mmHg per 90 days) and DBP (−0.66 vs −0.75 mmHg per 90 days) but steeper for RBS (−1.28 vs −0.77 mg/dL per 90 days) **(Supplementary Table 3)**. Restricting the sample to participants followed for more than 90 days changed the population-average SBP and DBP slopes little (−1.01 and −0.64 mmHg per 90 days) and attenuated the RBS slope from −1.28 to −0.94 mg/dL per 90 days **(Supplementary Table 4)**. The SBP and DBP slopes were robust to these choices; however, the RBS slope was more sensitive to the estimand and follow-up requirement.

Among groups classified by joint (HTN and DM) control status, SBP and DBP declined in the two groups with uncontrolled HTN and did not change significantly in the two groups with controlled HTN. RBS declined in the two groups with uncontrolled DM and increased in the two groups with controlled DM **(Supplementary Table 5)**. The largest RBS decline occurred among participants with controlled HTN and uncontrolled DM (−9.85 mg/dL per 90 days), compared with −5.82 mg/dL per 90 days among those uncontrolled for both conditions.

### 3.3 Odds of clinical control over time

The population-average odds of HTN control increased over follow-up (adjusted odds ratio [aOR] per 90 days: 1.21 [95% CI: 1.17, 1.25]), as did the odds of SBP control (1.22 [1.18, 1.27]) and DBP control (1.21 [1.16, 1.25]) **(Table 3)**. The odds of DM control increased modestly (1.06 [1.04, 1.08]). Unadjusted and adjusted ORs were nearly identical.

**Table 3:** Population-average odds of clinical control over follow-up.

| Outcome and model | OR per 30 days<br>(95% CI) | OR per 90 days<br>(95% CI) | OR per year<br>(95% CI) | p<br>value |
| --- | --- | --- | --- | --- |
| **HTN control** |  |  |  |  |
| Unadjusted | 1.06 (1.05, 1.08) | 1.21 (1.17, 1.25) | 2.14 (1.88, 2.44) | <0.001 |
| Adjusted | 1.06 (1.05, 1.08) | 1.21 (1.17, 1.25) | 2.15 (1.88, 2.45) | <0.001 |
| **SBP control** |  |  |  |  |
| Unadjusted | 1.07 (1.06, 1.08) | 1.22 (1.18, 1.27) | 2.26 (1.95, 2.62) | <0.001 |
| Adjusted | 1.07 (1.06, 1.08) | 1.22 (1.18, 1.27) | 2.26 (1.95, 2.63) | <0.001 |
| **DBP control** |  |  |  |  |
| Unadjusted | 1.06 (1.05, 1.08) | 1.20 (1.16, 1.25) | 2.11 (1.82, 2.45) | <0.001 |
| Adjusted | 1.06 (1.05, 1.08) | 1.21 (1.16, 1.25) | 2.14 (1.85, 2.48) | <0.001 |
| **DM control** |  |  |  |  |
| Unadjusted | 1.02 (1.01, 1.03) | 1.06 (1.04, 1.08) | 1.25 (1.16, 1.35) | <0.001 |
| Adjusted | 1.02 (1.01, 1.03) | 1.06 (1.04, 1.08) | 1.25 (1.16, 1.35) | <0.001 |

Across joint control groups, the odds of HTN control increased in the two groups with uncontrolled HTN at screening (aOR per 90 days: 1.45 [1.29, 1.63] and 1.39 [1.32, 1.46]) and did not change significantly in the two groups with controlled HTN **(Supplementary Table 6)**. The adjusted odds of DM control increased in the two groups with uncontrolled DM (1.13 [1.07, 1.20] and 1.29 [1.21, 1.37]) and decreased among participants with uncontrolled HTN and controlled DM (0.94 [0.90, 0.98]).

Within-person estimates led to the same conclusions, except that the decrease in the odds of DM control among participants controlled for both conditions was statistically significant only in the within-person model (0.92, p=0.028, vs 0.97, p=0.179).

### 3.4 Time to clinical control

The analytic sample for the exploratory time-to-control analysis included 754 participants with uncontrolled HTN at screening, of whom 624 achieved control during follow-up **(Figure 2A)**. Of the 636 with uncontrolled SBP, 538 achieved control **(Figure 2B)**. Of the 543 with uncontrolled DBP, 471 achieved control **(Figure 2C)**. Of the 494 with uncontrolled DM, 351 achieved control **(Figure 2D)**.

**Figure 2:**
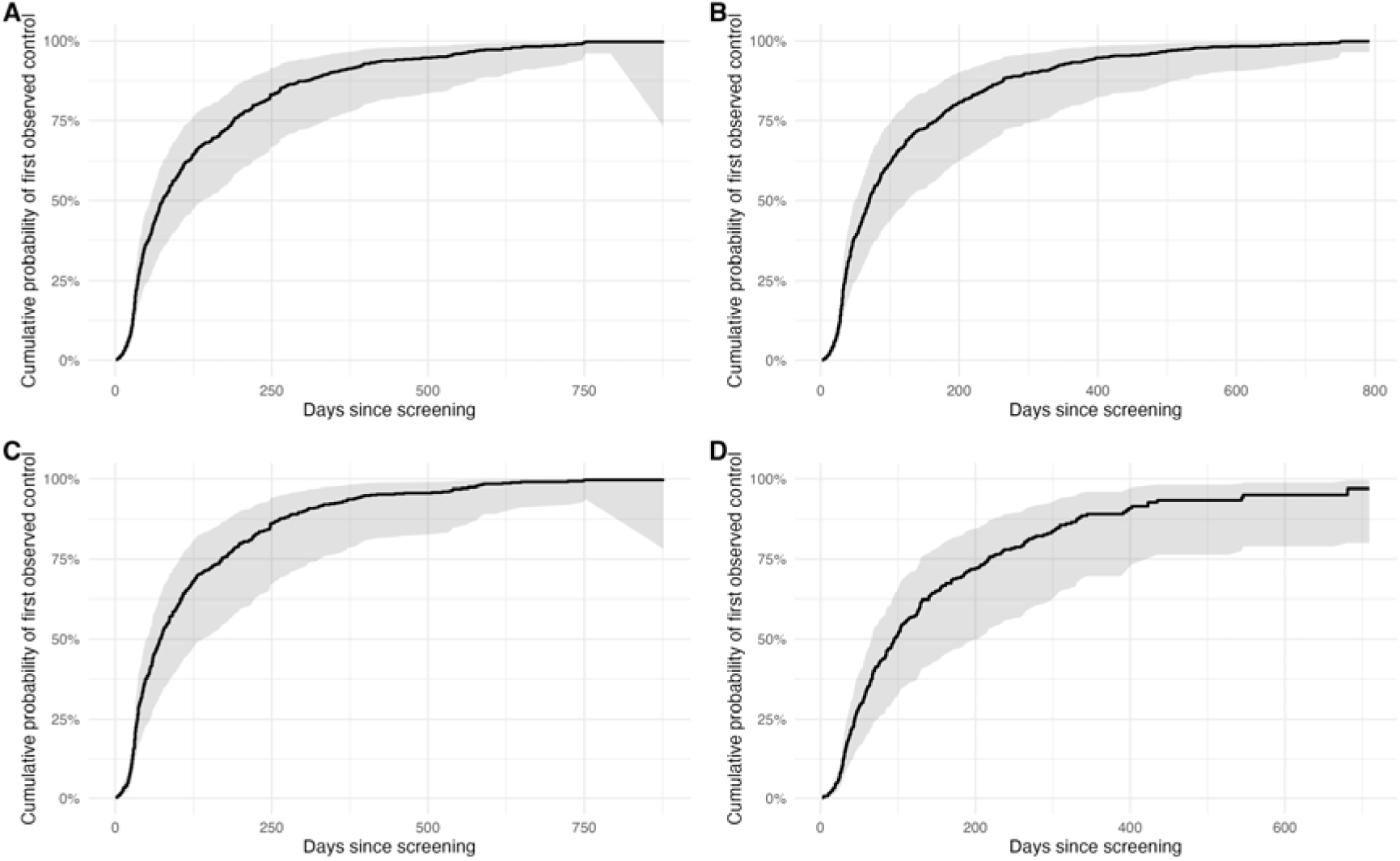
Cumulative probability of first observed control among participants uncontrolled at screening for (A) HTN control (n=754; 624 events), (B) SBP control (n=636; 538 events), (C) DBP control (n=543; 471 events), and (D) DM control (n=494; 351 events). Curves are model-based cumulative probabilities from the adjusted Cox models, evaluated at mean covariate values. Participants were drawn from those with more than one recorded visit before the 30-day follow-up restriction; time zero is screening. Shaded bands show 95% CIs. HTN: hypertension; SBP: systolic blood pressure; DBP: diastolic blood pressure; DM: diabetes mellitus; RBS: random blood sugar.

Global tests did not reject the proportional hazards assumption for HTN, SBP, and DM models (global p≥0.41). However, the assumption did not hold for the DBP model (p=0.028). The violation was driven by age (p=0.021). DBP hazard ratios, particularly for age, should be read as averages over follow-up.

Participants with controlled DM at screening reached HTN control more slowly than those with uncontrolled DM (hazard ratio [HR]: 0.68 [95% CI: 0.57, 0.81]). We found similar estimates for SBP control (0.67 [0.55, 0.81]) and DBP control (0.63 [0.51, 0.77]). Older age was associated with faster HTN control (HR per year: 1.010 [1.004, 1.017]), SBP control (1.008 [1.001, 1.015]), and DBP control (1.020 [1.012, 1.028]). No covariate was significantly associated with time to control for DM **(Supplementary Table 7)**.

## 4 Discussion

### 4.1 Summary of findings

This study examines changes in blood pressure and blood sugar among participants engaged with NEEM, a home-based primary care model for managing NCDs in rural India. Within participants, SBP declined by 1.19 mmHg, DBP by 0.75 mmHg, and RBS by 0.77 mg/dL per 90 days; the population-average odds of HTN and DM control increased by 21% and 6% per 90 days, respectively. Declines were concentrated among participants with uncontrolled HTN or DM at screening (SBP −2.43 mmHg and RBS −7.57 mg/dL per 90 days). Blood pressure did not change significantly among participants with controlled HTN at screening, whereas RBS rose slightly among those with controlled DM (1.33 mg/dL per 90 days). The change expected from regression to the mean was about half of the observed blood pressure decline and about two-thirds of the RBS decline between screening and the latest visit among participants uncontrolled at screening, so the blood pressure findings are more robust than the RBS findings.

These findings provide consistent evidence of improved HTN control and suggestive evidence of improved DM control among participants engaged in a digitally supported home-based primary care model in a rural Indian setting.

### 4.2 Public health and practice implications

Our results align with the broader shift toward integrated primary care models recommended by the WHO HEARTS and PEN frameworks. Previous IHCI evidence shows that blood pressure control rates were highest in primary healthcare facilities closer to patients.^11,13^ The NEEM program takes a further step along this continuum with a home-based, community health worker-led approach, supported by digital decision tools and medication delivery, to support NCD control among engaged participants.

Recent evidence further indicates that HTN is clustered in 14.9% of Indian households, yet these clusters account for 49.8% of all HTN cases in the country. Similarly, DM clustering in 7.7% of households contributes to 39.3% of the total DM burden.^26^ By targeting the household as the primary unit of intervention through regular home visits, the NEEM program could concentrate resources on households where these conditions cluster, potentially increasing the efficiency of screening and long-term interventions.

A comparable initiative, the Makkalai Thedi Maruthuvam program launched by the Government of Tamil Nadu, has shown similarly encouraging results for HTN and DM control.^27^ The clearer improvements in blood pressure than in blood sugar that we observed resemble the findings of a previous cluster randomized trial of community health worker home visits.^28^ A private, community health worker-led, technology-enabled program for DM and HTN among the urban poor in Hyderabad reported reductions in blood glucose and blood pressure among participants who remained enrolled, with 12-month retention of 16.5%.^29^ Like NEEM, such subscription-based private-sector models depend on sustained participant engagement.

The NEEM program brings together four elements: (i) home-based service delivery; (ii) protocol-driven care managed by locally recruited health workers assigned to defined patient groups; (iii) supportive supervision; and (iv) a subscription-based payment model covering medicines, home visits, and access to the care team. These elements were developed by drawing on evidence from successful health worker-led programs, such as the Alaska Community Health Aide Program, as well as insights from the IHCI.^11,30^ Notably, NEEM operates without a physical outpatient facility, an intentional design feature that reinforces home-based service culture. Health workers are employed full-time with compensation linked to the number of patients under their care. This model could bridge healthcare access gaps among underserved populations.

### 4.3 Strengths and limitations

This retrospective analysis of real-world programmatic data had a large sample size, repeated measurements within participants, and sensitivity analyses, which strengthened the interpretation of the findings.

However, there are several limitations. First, the study relied on routinely collected program data, which had high missingness for some predictors, including family size and use of DM and HTN medication. We could not include them as covariates in the analyses. Second, we assessed glycemic status using RBS, which varies with time since the last meal. Biomarkers such as HbA1c are better suited to evaluate DM control. DM control status may therefore have been misclassified. Third, visit timing was irregular and likely informative: participants with poor control, symptoms, or treatment changes may have returned at different intervals from those with stable values. GEE models account for within-person correlation but cannot address meaningful differences in visit timing. Fourth, the analyses relied on complete and plausible measurements. Therefore, selection bias may arise if missing or implausible values are related to participant characteristics or disease status. Results apply to participants with continued engagement (35% of those screened) and may not generalize to those with a single visit. Fifth, we lacked information on medication adherence, diet, physical activity, and biopsychosocial determinants, which can influence control rates. Finally, without a comparison group, the observed changes reflect the combined influence of care, medication changes, participant behavior, follow-up frequency, and regression to the mean. Hence, these findings are descriptive and should not be interpreted as causal effects of the program.

### 4.4 Future directions

Future research should assess long-term sustainability, cost-effectiveness, and patient experience. Evaluations that link process indicators to outcome measures and explore integration pathways into district health systems will be critical for scale-up. As India advances toward universal health coverage, hybrid home-based models such as NEEM could play a meaningful role in improving continuity and quality of chronic disease care.

The NEEM program aims to increase coverage and provide quality primary care through doorstep access and risk-based monitoring. The nominally priced, prepaid subscription model is also intended to make healthcare affordable and protect against catastrophic health expenditure. Regular home-based visits by trained locally recruited health workers are intended to support disease monitoring, improve treatment adherence, facilitate early intervention, and help overcome barriers such as access, affordability, and continuity of care.^11–13^ Studying the effects of such initiatives is needed to guide the development of effective healthcare delivery models to tackle the burgeoning burden of NCDs, particularly in the private sector.

## 5 Conclusion

The rising burden of non-communicable diseases in India and the large number of individuals without health insurance necessitate an innovative and scalable service delivery model. Our primary and sensitivity analyses provide consistent evidence of improved HTN control and suggestive evidence of improved DM control among those engaged in the NEEM program, a home-based, digitally enabled, local health worker-led primary care model in rural Maharashtra, India. As India strives toward universal health coverage, structured home-based primary care built on early screening and protocol-driven interventions delivered by community health workers may offer a practical approach to reaching underserved rural populations.

## Supporting information

Appendix with Supplementary Tables and Figure

STROBE Checklist

## Data Availability

All data produced in the present study are available upon reasonable request to the authors

## Declarations

## Ethics approval and consent to participate

The study was approved by the Indian Association for Preventive and Social Medicine Ethics Committee (IAPSM/00112024). Digital written informed consent was obtained from all participants.

## Consent for publication

Not applicable. This manuscript does not contain any individual person’s data in any form (including any individual details, images, or videos). All data presented are in aggregate form.

## Availability of data and materials

The datasets used and/or analyzed in the current study are available from the corresponding author upon reasonable request, after approval from the relevant ethics committee.

## AI use disclosure

We used Claude Opus 5.5 (Cowork) for code validation, annotation, and text checking and editing for language, and Grammarly AI for proofreading. AI tools did not have access to identifiable individual participant data.

## Authors’ contributions

UG, LK, and SZ conceptualized the study. SZ and JSM performed the data analysis and visualization. SZ interpreted the results. UG and LK wrote the initial draft of the manuscript. All authors reviewed, edited, and approved the final manuscript.

## Acknowledgments

We thank Bindu Ananth, Shreesha B V, and Kruthi N for their help with the study, and everyone else at ASAR for their support.

