## Appendix with Supplementary Tables and Figure for "Home-based primary care services for managing non-communicable diseases in rural India: a retrospective multiple time-point observational study"

Contents

Supplementary Table 1: Physiologic plausibility ranges and values set to missing

Supplementary Table 2: Regression-to-the-mean diagnostics by screening control status

Supplementary Table 3: Within-person versus population-average adjusted time slopes

Supplementary Table 4: Sensitivity of population-average slopes to a longer follow-up requirement

Supplementary Table 5: Change in SBP, DBP, and RBS by joint HTN and DM control status at screening

Supplementary Table 6: Odds of HTN and DM control by joint control status at screening

Supplementary Table 7: Cox models: factors associated with time to first observed control

Supplementary Figure 1: Village-level distribution of the primary analytic sample, Satara district, Maharashtra, India

Abbreviations: BP: blood pressure; SBP: systolic blood pressure; DBP: diastolic blood pressure; RBS: random blood sugar; HTN: hypertension; DM: diabetes mellitus; CI: confidence interval; SE: standard error; OR: odds ratio; HR: hazard ratio; RTM: regression to the mean.

**Supplementary Table 1: Physiologic plausibility ranges and values set to missing.**

| **Measurement** | **Plausible range** | **Values set to missing** |
| --- | --- | --- |
| Height | 121–188 cm | 69 |
| Weight | 25–125 kg | 38 |
| Oxygen saturation (SpO₂) | 70–100% | 150 |
| Pulse rate | 50–120 beats/min | 185 |
| Respiratory rate | 10–24 breaths/min | 102 |
| Body temperature | 90–110 °F | 321 |
| Waist circumference | 50.8–152.4 cm | 8 |
| RBS at screening | 20–600 mg/dL | 37 |
| RBS at follow-up visits | 20–600 mg/dL | 830 |
| SBP at screening | 60–300 mmHg | 0 |
| SBP at follow-up visits | 60–300 mmHg | 15 |
| DBP at screening | 40–200 mmHg | 0 |
| DBP at follow-up visits | 40–200 mmHg | 18 |
| RBS at latest visit | 20–600 mg/dL | 346 |
| SBP at latest visit | 60–300 mmHg | 9 |
| DBP at latest visit | 40–200 mmHg | 2 |

Ranges were applied to the 15,233 observations of participants with more than one visit and more than 30 days of follow-up. Values outside the range were set to missing; no visit was removed at this step, so the number of observations was unchanged. Control status at screening, at each follow-up visit, and at the latest visit was then derived from the cleaned values. Counts are visit-level, so latest-visit values repeated across a participant’s visits are counted once per visit. The analytic sample for the exploratory time-to-control analysis (15,757 observations, drawn before the 30-day restriction) was cleaned with the same ranges; because it is larger, its counts of values set to missing are slightly higher.

**Supplementary Table 2: Regression-to-the-mean diagnostics by screening control status.**

| **Stratum at screening** | **N** | **Mean at screening** | **Mean at latest visit** | **Observed change** | **Change expected from RTM** | **Change beyond RTM** | **Correlation r** |
| --- | --- | --- | --- | --- | --- | --- | --- |
| **SBP, mmHg (strata by SBP control)** | | | | | | | |
| Uncontrolled | 607 | 158.6 | 136.4 | −22.17 | −12.63 | −9.54 | 0.403 |
| Controlled | 1,061 | 125.3 | 127.0 | 1.69 | 7.22 | −5.53 | 0.403 |
| **SBP, mmHg (strata by HTN control)** | | | | | | | |
| Uncontrolled | 719 | 154.4 | 135.5 | −18.96 | −10.16 | −8.80 | 0.403 |
| Controlled | 949 | 124.5 | 126.6 | 2.08 | 7.70 | −5.62 | 0.403 |
| **DBP, mmHg (strata by DBP control)** | | | | | | | |
| Uncontrolled | 515 | 99.1 | 85.4 | −13.66 | −8.38 | −5.28 | 0.394 |
| Controlled | 1,153 | 79.0 | 79.5 | 0.49 | 3.74 | −3.25 | 0.394 |
| **DBP, mmHg (strata by HTN control)** | | | | | | | |
| Uncontrolled | 719 | 94.1 | 83.8 | −10.26 | −5.36 | −4.90 | 0.394 |
| Controlled | 949 | 78.5 | 79.5 | 0.96 | 4.06 | −3.10 | 0.394 |
| **RBS, mg/dL (strata by DM control)** | | | | | | | |
| Uncontrolled | 451 | 303.4 | 227.3 | −76.10 | −51.78 | −24.32 | 0.598 |
| Controlled | 1,096 | 121.7 | 133.3 | 11.66 | 21.31 | −9.65 | 0.598 |

Values are each participant’s screening and latest-visit values. The change expected from RTM is approximately (r − 1) × (stratum mean at screening − overall mean at screening), where r is the correlation between participants’ screening and latest values. Because r conflates true change with measurement noise, it tends to understate reliability and overstate RTM, so the change beyond RTM is a conservative estimate. Pure RTM moves uncontrolled and controlled strata in opposite directions; a change beyond RTM with the same sign in both strata indicates change in addition to mean reversion. Each row includes participants with both screening and latest values, so the RBS rows (1,547 participants) exclude those without a latest RBS.

**Supplementary Table 3: Within-person versus population-average adjusted time slopes.**

| **Outcome and estimand** | **Daily coefficient (SE)** | **Change per 90 days** | **Projected annual change (95% CI)** | **p value** |
| --- | --- | --- | --- | --- |
| **SBP, mmHg** | | | | |
| Within-person (primary) | −0.0133 (0.0010) | −1.19 | −4.84 (−5.58, −4.10) | <0.001 |
| Population-average | −0.0118 (0.0009) | −1.06 | −4.31 (−4.96, −3.66) | <0.001 |
| **DBP, mmHg** | | | | |
| Within-person (primary) | −0.0083 (0.0006) | −0.75 | −3.04 (−3.49, −2.58) | <0.001 |
| Population-average | −0.0073 (0.0006) | −0.66 | −2.67 (−3.07, −2.27) | <0.001 |
| **RBS, mg/dL** | | | | |
| Within-person (primary) | −0.0085 (0.0036) | −0.77 | −3.12 (−5.66, −0.57) | 0.016 |
| Population-average | −0.0142 (0.0033) | −1.28 | −5.19 (−7.55, −2.83) | <0.001 |

Adjusted models (Model 2), primary analytic sample (1,668 participants; 12,769 observations). Within-person slopes are the coefficient on the within-person deviation of follow-up time from the within-between (Mundlak) decomposition; population-average slopes are from the corresponding standard generalized estimating equation. Projected annual change is a linear projection (daily coefficient × 365) that may extend beyond the observed follow-up range.

**Supplementary Table 4: Sensitivity of population-average slopes to a longer follow-up requirement.**

| **Outcome and sample** | **Change per 90 days (95% CI)** | **p value** |
| --- | --- | --- |
| **SBP, mmHg** | | |
| More than 30 days of follow-up (primary analytic sample) | −1.06 (−1.22, −0.90) | <0.001 |
| More than 90 days of follow-up | −1.01 (−1.17, −0.85) | <0.001 |
| **DBP, mmHg** | | |
| More than 30 days of follow-up (primary analytic sample) | −0.66 (−0.76, −0.56) | <0.001 |
| More than 90 days of follow-up | −0.64 (−0.74, −0.54) | <0.001 |
| **RBS, mg/dL** | | |
| More than 30 days of follow-up (primary analytic sample) | −1.28 (−1.86, −0.70) | <0.001 |
| More than 90 days of follow-up | −0.94 (−1.53, −0.35) | 0.002 |

Adjusted population-average generalized estimating equation slopes (change per 90 days = daily coefficient × 90). The more-than-90-day models used outcome-specific complete cases (BP models did not require a follow-up RBS value), whereas the primary analytic sample required complete data on all primary model variables.

**Supplementary Table 5: Change in SBP, DBP, and RBS by joint HTN and DM control status at screening.**

| **Joint control status at screening** | **Participants (observations)** | **Within-person slope per 90 days (95% CI)** | **p value** | **Population-average slope per 90 days (95% CI)** | **p value** |
| --- | --- | --- | --- | --- | --- |
| **SBP, mmHg per 90 days (omnibus p<0.001)** | | | | | |
| Both uncontrolled | 196 (1,729) | −2.25 (−2.81, −1.70) | <0.001 | −2.09 (−2.58, −1.60) | <0.001 |
| HTN controlled, DM uncontrolled | 256 (1,801) | 0.15 (−0.25, 0.55) | 0.466 | 0.24 (−0.12, 0.60) | 0.193 |
| HTN uncontrolled, DM controlled | 523 (4,229) | −2.49 (−2.81, −2.18) | <0.001 | −2.28 (−2.56, −2.00) | <0.001 |
| Both controlled | 693 (5,010) | 0.09 (−0.09, 0.26) | 0.325 | 0.06 (−0.11, 0.23) | 0.477 |
| **DBP, mmHg per 90 days (omnibus p<0.001)** | | | | | |
| Both uncontrolled | 196 (1,729) | −1.21 (−1.48, −0.95) | <0.001 | −1.19 (−1.44, −0.93) | <0.001 |
| HTN controlled, DM uncontrolled | 256 (1,801) | −0.04 (−0.29, 0.21) | 0.760 | 0.00 (−0.22, 0.22) | 0.985 |
| HTN uncontrolled, DM controlled | 523 (4,229) | −1.48 (−1.70, −1.27) | <0.001 | −1.32 (−1.50, −1.14) | <0.001 |
| Both controlled | 693 (5,010) | −0.06 (−0.18, 0.07) | 0.370 | −0.05 (−0.17, 0.07) | 0.409 |
| **RBS, mg/dL per 90 days (omnibus p<0.001)** | | | | | |
| Both uncontrolled | 196 (1,729) | −5.82 (−8.44, −3.19) | <0.001 | −5.67 (−8.22, −3.11) | <0.001 |
| HTN controlled, DM uncontrolled | 256 (1,801) | −9.85 (−12.95, −6.76) | <0.001 | −9.95 (−12.68, −7.22) | <0.001 |
| HTN uncontrolled, DM controlled | 523 (4,229) | 1.59 (0.94, 2.24) | <0.001 | 1.14 (0.52, 1.76) | <0.001 |
| Both controlled | 693 (5,010) | 1.09 (0.49, 1.69) | <0.001 | 0.52 (−0.02, 1.05) | 0.058 |

One generalized estimating equation per outcome with follow-up time interacted with the four-group joint control variable; screening control covariates were omitted from the adjustment set because the group encodes them. Omnibus p is the joint Wald test that group trajectories differ (within-person model). 95% CIs and p values use the robust sandwich variance. Groups are defined on single screening measurements and are subject to regression to the mean.

**Supplementary Table 6: Odds of HTN and DM control by joint control status at screening.**

| **Joint control status at screening** | **Population-average OR per 90 days (95% CI)** | **p value** | **Within-person OR per 90 days (95% CI)** | **p value** |
| --- | --- | --- | --- | --- |
| **HTN control (omnibus p<0.001)** | | | | |
| Both uncontrolled | 1.45 (1.29, 1.63) | <0.001 | 1.50 (1.36, 1.64) | <0.001 |
| HTN controlled, DM uncontrolled | 0.99 (0.89, 1.11) | 0.886 | 1.05 (0.92, 1.19) | 0.489 |
| HTN uncontrolled, DM controlled | 1.39 (1.32, 1.46) | <0.001 | 1.46 (1.40, 1.53) | <0.001 |
| Both controlled | 0.98 (0.93, 1.02) | 0.324 | 0.99 (0.94, 1.05) | 0.807 |
| **DM control (omnibus p<0.001)** | | | | |
| Both uncontrolled | 1.13 (1.07, 1.20) | <0.001 | 1.14 (1.07, 1.22) | <0.001 |
| HTN controlled, DM uncontrolled | 1.29 (1.21, 1.37) | <0.001 | 1.28 (1.19, 1.38) | <0.001 |
| HTN uncontrolled, DM controlled | 0.94 (0.90, 0.98) | 0.002 | 0.88 (0.82, 0.95) | <0.001 |
| Both controlled | 0.97 (0.93, 1.01) | 0.179 | 0.92 (0.85, 0.99) | 0.028 |

Group sizes are as in Supplementary Table 5. Population-average ORs are presented first, consistent with the main analysis of control, because the within-between decomposition is only approximate in logistic models. Omnibus p values from the population-average and within-person models were both <0.001. 95% CIs and p values use the robust sandwich variance.

**Supplementary Table 7: Cox models: factors associated with time to first observed control.**

| **Characteristic** | **HR** | **95% CI** | **p value** |
| --- | --- | --- | --- |
| **HTN control (n=754; 624 events)** | | | |
| Age, per year | 1.01 | 1.00, 1.02 | 0.002 |
| Sex: male (ref. female) | 0.95 | 0.81, 1.11 | 0.507 |
| Smoking: former smoker (ref. current smoker) | 0.56 | 0.26, 1.21 | 0.139 |
| Smoking: non-smoker (ref. current smoker) | 1.18 | 0.84, 1.67 | 0.343 |
| Known chronic kidney disease (ref. no) | Not estimableᵃ | — | — |
| Known cardiovascular disease (ref. no) | 0.85 | 0.64, 1.12 | 0.244 |
| DM controlled at screening (ref. uncontrolled) | 0.68 | 0.57, 0.81 | <0.001 |
| **SBP control (n=636; 538 events)** | | | |
| Age, per year | 1.01 | 1.00, 1.02 | 0.030 |
| Sex: male (ref. female) | 0.97 | 0.81, 1.15 | 0.689 |
| Smoking: former smoker (ref. current smoker) | 0.54 | 0.24, 1.23 | 0.144 |
| Smoking: non-smoker (ref. current smoker) | 1.14 | 0.79, 1.65 | 0.476 |
| Known chronic kidney disease (ref. no) | Not estimableᵃ | — | — |
| Known cardiovascular disease (ref. no) | 0.92 | 0.70, 1.22 | 0.554 |
| DM controlled at screening (ref. uncontrolled) | 0.67 | 0.55, 0.81 | <0.001 |
| **DBP control (n=543; 471 events)** | | | |
| Age, per year | 1.02 | 1.01, 1.03 | <0.001 |
| Sex: male (ref. female) | 1.06 | 0.88, 1.27 | 0.569 |
| Smoking: former smoker (ref. current smoker) | 0.87 | 0.30, 2.54 | 0.805 |
| Smoking: non-smoker (ref. current smoker) | 1.28 | 0.87, 1.89 | 0.212 |
| Known chronic kidney disease (ref. no) | Not estimableᵃ | — | — |
| Known cardiovascular disease (ref. no) | 0.73 | 0.52, 1.03 | 0.077 |
| DM controlled at screening (ref. uncontrolled) | 0.63 | 0.51, 0.77 | <0.001 |
| **DM control (n=494; 351 events)** | | | |
| Age, per year | 1.00 | 0.99, 1.01 | 0.882 |
| Sex: male (ref. female) | 1.01 | 0.81, 1.26 | 0.935 |
| Smoking: former smoker (ref. current smoker) | 1.48 | 0.62, 3.51 | 0.375 |
| Smoking: non-smoker (ref. current smoker) | 1.01 | 0.67, 1.55 | 0.948 |
| Known chronic kidney disease (ref. no) | 1.31 | 0.61, 2.83 | 0.494 |
| Known cardiovascular disease (ref. no) | 1.05 | 0.66, 1.65 | 0.844 |
| HTN controlled at screening (ref. uncontrolled) | 1.16 | 0.94, 1.45 | 0.174 |

Analytic sample for the exploratory time-to-control analysis: participants uncontrolled at screening for each outcome, drawn from those with more than one recorded visit before the 30-day follow-up restriction. HR >1 indicates faster first observed control. Time zero is screening; participants in whom control was not observed were censored at their last observed follow-up measurement.

ᵃ The model did not produce a finite estimate for known chronic kidney disease in the HTN, SBP, and DBP control models (the model returned HR 0.00 with 95% CI 0.00 to infinity).

**Supplementary Figure 1: Village-level distribution of the primary analytic sample, Satara district, Maharashtra, India, showing (A) participants, (B) visits, (C) the percentage with HTN controlled at screening, and (D) the percentage with DM controlled at screening, across villages.**

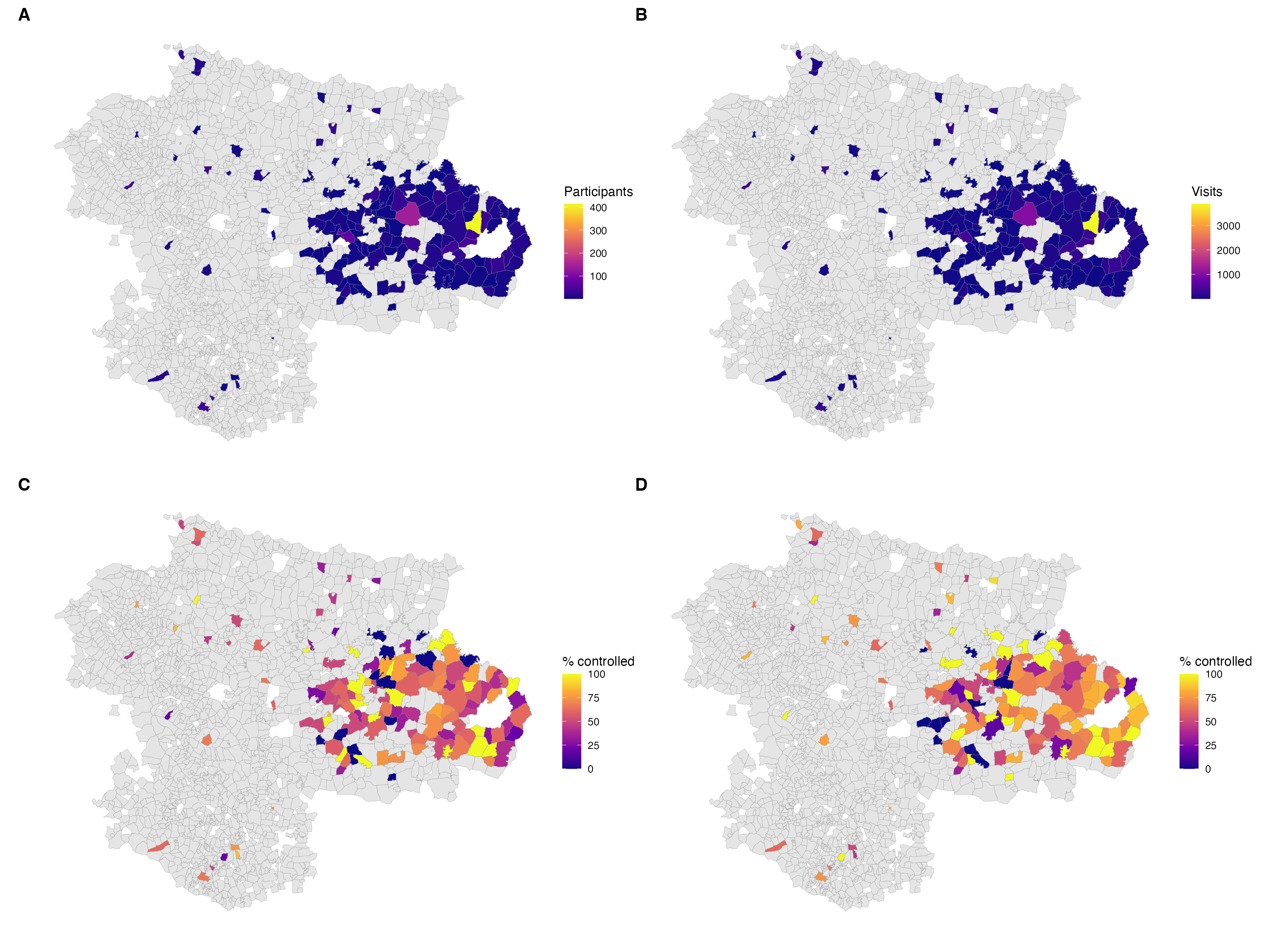

Values are for the primary analytic sample; villages in grey had no participants. Village boundaries are from the 2011 Census. Nine villages (173 participants) were outside the shapefile and are not shown.
